## Supplementary methods, figures and tables for "Viral kinetic modeling and clinical trial simulation predicts disruption of respiratory disease trials by non-pharmaceutical COVID-19 interventions"

#### ABSTRACT

Clinical research in infectious respiratory diseases has been profoundly affected by non-pharmaceutical interventions (NPIs) against COVID-19. On top of trial delays or even discontinuation which have been observed in all disease areas, NPIs altered transmission pattern of many seasonal respiratory viruses which followed regular patterns for decades before the pandemic. Clinical trial design based on pre-pandemic historical data therefore needs to be put in question. In this article, we show how knowledge-based mathematical modeling can be used to address this issue. We set up an epidemiological model of respiratory tract infection (RTI) sensitive to a time dependent between-host transmission rate and coupled this model to a mechanistic description of viral RTI episodes in an individual patient. By reducing the transmission rate when the lockdown was introduced in the United Kingdom in March 2020, we were able to reproduce the perturbed 2020 RTI disease burden data. Using this setup, we simulated several NPIs scenarios of various strength (none, mild, medium, strong) and conducted placebo-controlled *in silico* clinical trials in pediatric patients with recurrent RTIs (RRTI) quantifying annual RTI rate distributions. In interventional arms, virtual patients aged 1-5 years received the bacterial lysate OM-85 (approved in several countries for the prevention of pediatric RRTIs) through a pro-type I immunomodulation mechanism of action described by a physiologically based pharmacokinetics and pharmacodynamics approach (PBPK/PD). Our predictions showed that sample size estimates based on the ratio of RTI rates (or the post-hoc power of fixed sample size trials) are not majorly impacted under NPIs which are less severe (none, mild and medium NPIs) than a strict lockdown (strong NPI). However, NPIs show a stronger impact on metrics more relevant for assessing the clinical relevance of the effect such as absolute benefit. This dichotomy shows the risk that successful trials (even with their primary endpoints being met) still get challenged in risk benefit assessment during the review of market authorization. Furthermore, we found that a mild NPI scenario already affected the time to recruit significantly when sticking to eligibility criteria complying with historical data. In summary, our model predictions can help rationalize and forecast post-COVID-19 trial feasibility. They advocate for gauging absolute and relative benefit metrics as well as clinical relevance for assessing efficacy hypotheses in trial design and they question eligibility criteria misaligned with the actual disease burden.

#### Supplementary Methods

##### Within-host viral infection disease model

To date, several modeling efforts have succeeded to model viral kinetics during a respiratory tract infection (RTI) mainly for influenza[1,2]. Building on this, we developed a disease model which attempts to capture the interplay between viral infections and immune dynamics in the respiratory tract tissues. The model accounts for the following mechanisms: 1) Exposure to the pathogen, onset and resolution of infection; 2) Target cell infection and pathogen replication; 3) Preemptive clearance by the innate immune system; 4) Innate inflammatory response; 5) Activation of the adaptive response; 6) Neutralization of the virus by antibodies; 7) Cytolysis of infected cells by cytotoxic lymphocytes; 8) Age-dependent maturation of the immune system; 9) General immunocompetence. The resulting ODE system is detailed as follows.

##### *Variables*

$E_h$ Healthy epithelial cell
$E_i$ Infected epithelial cell
$Ig$ Virus-specific immunoglobulins produced by B cells during infection
$Ig_A$ Non-specific immunoglobulins produced by OM-85 induced plasma cells
$L$ Cytotoxic T lymphocytes
$V$ Virions
$M$ Pre-activated type 1 innate cells

#### ODEs

$$\begin{aligned}
\dot{E}_h =& p_h \cdot \bar{E}_h \left(1 - \frac{E_h + E_i}{\frac{\bar{E}_h \cdot p_h}{p_h - d_h}}\right) - d_h E_h - k_{inf} \cdot v_f E_h V \frac{1}{\left(1+\frac{c}{V}\right)^2} \\
\dot{E}_i =& k_{inf} \cdot v_f E_h V \frac{1}{\left(1+\frac{c}{V}\right)^2} - d_i E_i - n_i L E_i \\
\dot{Ig} =& p_{ig} \cdot S \frac{V}{K_V + V}\left(1 + p_M \frac{M}{K_M + M}\right) - d_{ig} Ig \\
\dot{Ig}_A =& - d_{IgA} \cdot Ig_A \\
\dot{L} =& - d_L L + p_L \cdot S \frac{V}{K_V + V}\left(1 + p_M \frac{M}{K_M + M}\right) \\
\dot{V} =& - k_{inf} \cdot v_f E_h V \frac{1}{\left(1+\frac{c}{V}\right)^2} + p_v \cdot v_f E_i \frac{1}{\left(1+\frac{c}{E_i}\right)^2} - d_v V - n_v \left(Ig + Ig_A\right) V
\end{aligned}$$

#### Parameters

In the context of a single RTI, we calibrated the set of unknown parameters of the model against mean viral load of individual profiles obtained during an experimental human RSV (HRSV) infection[3] (Figure S2). The complete set of parameters is described in Table S1.

#### Between-host SIRS model

The between-host SIRS model follows the classical formalism[4,5] with populations of size $N$, partitioned into susceptible $S$, infected $I$ and recovered $R$ populations. Healthy susceptible individuals $S$ may be infected with an infection rate $\beta$, leading them to become infected individuals $I$. Those can then recover after a given time with a recovery rate $\gamma$, leading them to become recovered individuals $R$. Since we are interested in recurrent infections, recovered individuals $R$ may lose their immunity after a given time, represented by an immunity loss rate $\zeta$, and become as a consequence susceptible again ($S$) to an infection. The system of differential equations reads:

$$\begin{aligned}
\dot{S} &= -\beta \cdot S \cdot I + \zeta \cdot R \\
\dot{I} &= \beta \cdot S \cdot I - \gamma \cdot I \\
\dot{R} &= \gamma \cdot I - \zeta \cdot R
\end{aligned}$$

The most common respiratory viruses causing RTIs worldwide in children are respiratory syncytial viruses (RSV), influenza viruses and rhinoviruses[6,7]. Therefore, we used three parallel SIRS models for each virus class without discriminating between viral strain. To capture seasonal variation of temperature in temperate climates[8], we used the common approach[9] where a periodic time-varying function is considered instead of a constant taken for the transmission parameter $\beta$: $\beta(t) = b_0 \cdot (1 + b_1 \cdot cos(2\pi t + \phi))$. For rhinoviruses, which show approximately a biannual behaviour[10], we divided the period by two.

The virus-dependent sets of parameters were calibrated using virus-specific incidence data[11–13] (Figure S4) and are detailed in Table S2. Furthermore, each of the respiratory viruses has a unique susceptibility to cause either an upper RTI (URTI) or lower RTI (LRTI)[14,15], which we use for probabilistic guidance on the determination of whether an RTI is a normally milder URTI or a usually more severe LRTI[16].

To include the impact of COVID-19 containment measures, we simply decrease the transmission parameter $\beta$ similarly to what is often reported in the literature[17,18] (Figure S1). This allows to explore the impact of the strength of the containment measures.

#### PBPK/PD model of OM-85 effect

OM-85 is administered orally and is absorbed in the intestine, triggering gut-associated lymphoid tissue (GALT) stimulation and subsequently generating the immune response within mucosal tissue (MALT) in other organs such as the respiratory tract[19]. The key factors in this chain-like reaction are reactive Peyers Patches (PPs) of GALT, responsible for antigen identification and subsequent generation of the adequate response. To model this, we used a physiologically based pharmacokinetic (PBPK) model coupled with a pharmacodynamics (PD) model of immune response in the GALT PPs. PBPK models map the complex drug transport scheme onto a physiologically realistic compartmental structure and in this sense are predetermined and largely independent of the particular drug of interest[20]. We incorporated a gut absorption model (Advanced CAT model[21]) but disregarded disintegration and dissolution for simplicity since the bacterial lysates such as OM-85 are highly soluble in water. The PBPK structure of the model allows us to use systemic blood circulation and lymphatic circuits to describe migration of activated immune cells from Peyer's Patches to mesenteric lymph nodes into the bloodstream with consecutive homing to effector sites such as respiratory tract mucosal tissue (Figure S5).

Pharmacokinetics data are available for a radiolabelled derivative of a product (OM-89) similar to OM-85 but only in rodents[22,23]. To incorporate those and parameterize the PK drug-specific parameters, we transformed our PBPK model into an inter-species model by allometric scaling and species-specific parameters[24–29]. Based on these data (25% of radioactivity detected in exhaled air) we incorporated a non-specific non-renal clearance as well as a metabolic clearance in the liver. Result of this calibration are presented in Figure S6 for agreement between the data and the simulations and in Table S3 for the values of the calibrated parameters.

For the pharmacodynamics effect of OM-85 in the GALT: in the first stage, when OM-85 reaches PPs, interactions between pathogen-associated molecular pattern (PAMPs) and pattern recognition receptors (PRR) induce a nonspecific activation of innate immune cells, including macrophage, monocyte, dendritic cell, natural killer cells and granulocyte with cytokines and chemokines production, activation of phagocytosis and early pathogens destruction[30,31]. In the second stage, antigen-specific T and B cells are generated in the PPs as well as a considerable number of lymphoblasts, mostly immunoglobulin A (IgA) precursors of the IgA producing plasmocytes[31]. In a last stage, lymphocytes and lymphoblasts maturate in mesentertic lymph nodes (mLNs) and subsequently migrate into MALT of various organs[31], leading to an increased production of antibacterial antibodies in serum, saliva, PPs, mLNs, but also the respiratory tract[30]. In order to model these mechanisms, we built a model that focuses on the perturbation of GALT homeostasis resulting from OM-85 administration in the Peyer's patches (Figure S5). Because of the scarcity of pharmacodynamic data and markers in human, we did not attempt to represent the complex interactions between all immune cells in GALT at homeostasis and instead focus on a few key immune cells and interactions in the immunomodulatory response to OM-85 (dendritic cells, innate cells, IgA+ B and plasma cells, and regulatory T cells). The resulting ODE system (in two representation compartments: a Peyer's Patch and the respiratory tract tissue) is presented below.

##### *Variables*

$D$ Dendritic cells
$O$ OM-85 drug
$M_p$ Reprogrammed type-1 innate progenitors
$M$ Pre-activated type 1 innate cells
$B_L$ IgA+ memory B cells
$B_P$ IgA+ plasmacells
$T_r$ Regulatory T cells
$Ig_A$ Non-specific IgA
$X_V$ Concentration of $X$ in the vascular compartment of a given organ

##### *ODEs*

**Peyer's Patch**

$$\dot{D} = E_O \cdot \frac{O^h}{K_O^h + O^h} - d_D \cdot D$$

$$\dot{M_p} = E_{M_p} \frac{D}{K_{M_p} + D} - (\alpha + d_{M_p}) \cdot M_p$$

$$\dot{M} = \alpha \cdot M_p - L_{PP} \cdot (1 - \sigma_{PP}) \cdot M$$

$$\dot{B_L} = E_{B_L} \frac{D}{K_{B_L} + D} - (\beta + d_{B_L}) \cdot B_L$$

$$\dot{B_P} = \left( \beta + E_{B_P} \frac{D}{K_{B_P} + D} \right) B_L - (d_{B_P} + L_{PP} \cdot (1 - \sigma_{PP})) \cdot B_P$$

$$\dot{T_r} = E_{T_r} \frac{D}{K_{T_r} + D} - (d_{T_r} + L_{PP} \cdot (1 - \sigma_{PP})) \cdot T_r$$

###### Respiratory tract tissue

$$\dot{M} = L \cdot \sigma_V \cdot (1 - \sigma_V^S) \cdot M_V - d_M \cdot M$$
$$\dot{B_P} = L \cdot \sigma_V \cdot (1 - \sigma_V^S) \cdot B_{LV} - d_{B_P} \cdot B_P$$
$$\dot{T_r} = L \cdot \sigma_V \cdot (1 - \sigma_V^S) \cdot T_{rV} - d_{T_r} \cdot T_r$$
$$\dot{Ig_A} = p_{Ig_A} + \gamma \cdot B_P - d_{Ig_A} \cdot Ig_A$$

We calibrated the model using data reported by Lusuardi *et al.* (1993) who studied IgA levels in bronchoalveolar lavage fluids (BAL) after treatment with OM-85 (Figure S7, Table S4). Note that this represent very few data points compared to the number of degrees of freedom of our model. This is why, we validated this calibration using data reported by Danek et al. (1996)[32] who used a treatment regimen different from the one in Lusuardi *et al.* (1993) (Figure S8).

#### Impact of age

The immune system evolves throughout life with major differences between infants, young children, adults and elderly and as such, age is a determinant factor for susceptibility to respiratory tract infections (and potentially response to OM-85 treatment). The increased susceptibility of infants and young children to respiratory infections is the result of the physiological immaturity of components of the systemic and local immune responses and, possibly, of suboptimal complex crosstalk between microbiota and immune system effectors[33]. For instance, neutrophils from neonates exhibit defective bactericidal activity and the lower DC efficiency contributes to sustain the early at birth Th2 bias, related to elevated intrauterine IL-4 and IL-10 production[33,34].

To account for this immune system maturation, we implemented an age-dependent modulation of a subset of immune-related parameters (e.g. $E_O$, $p_L$) of the following form:

$$p(x) = p^* \cdot \frac{1 + \alpha \cdot e^{-k(x-a)}}{1 + e^{-k(x-a)}}$$

$p(x)$ Parameter value as a function of age in years
$x$ Age in years
$p^*$ Reference parameter value for an adult
$\alpha$ Maximum (relative) decrease of parameter value due to immature immune system
$k$ Immune maturation slope
$a$ Immune maturation inflection point

The set of impacted parameters includes: $E_O$ for activation of DCs by OM-85, $p_L$ for recruitment of virus-specific T cells and $p_{ig}$ for production of virus-specific immunoglobulins. We calibrated the parameters controlling age-dependency in order to reproduce the age-dependent distribution of number of RTIs observed in the COPSAC2000 birth cohort[35] (Figure S3) and obtained $k = 2$ and $a = 3$ years for the three impacted parameters and $\alpha = 0.5$ for DC activation and $\alpha = 0.85$ for the two other mechanisms.

##### Interface of between host and within host model of viral infection

The time-dependent solution of the SIRS model corresponds to the instantaneous prevalence of RTI caused by any of the described viruses. We utilize the time-modulation of the prevalence to model the probability density $p(t)$ of getting exposed to an RTI provoking virus at time $t$ for a representative (mean) individual patient. We assume that any number of RTIs needs an equal or bigger number of exposures to viruses causing the infection and that the intrinsic properties of a patient (notably his immune system) will determine how many of these exposures will lead to a detectable RTI. First, we follow a Monte Carlo-like process to determine potential exposure time points during the simulation (24 bins per year, amounting to maximum of 48 potential exposures over 2 years simulation). We define the acceptance criterion $W(ex)$ as:

$$W(ex) = \begin{cases} True \text{ if } ex \leq p(t) \\ False \text{ if } ex > p(t) \end{cases} \quad (1)$$

We denote $ex$ to be the potential exposure, which is randomly drawn from a (scalable) uniform distribution $U_{[0,\gamma]}$, and is compared to the instantaneous probability density of exposure in each bin. An individual is exposed to a respiratory virus if the acceptance criterion (1) is evaluated as *True*. Following this procedure, the distribution of the number of exposures in a given time period $[t_0, t_A]$ resembles a (truncated) Poisson distribution with parameter $\lambda$.

The deterministic immunological model transforms exposures into RTIs and the value of $\gamma$ can be adapted so that the combined deterministic-stochastic model can reproduce the expected mean number of RTIs in a specific period for a reference population. The number of RTIs for individual patients and subgroups will also depend on the state of their immune system. In the within-host model, the individual immune system states of patients are characterised by inter-patient and inter-observation variability, both following a random statistical model around an average value and consequently, there are additional parameters determining the fate of each individual exposure described above. In fact, the mean and variance of the distribution of the immune system's state related parameters determine the transformation function between the distribution of exposure (Poisson-like) counts and the distribution of RTI counts, so that we conveniently fix $\gamma$ to a number of 10 (large enough to encompass a wide range of behaviors in terms of number of RTIs per year), and calibrate only the random immune system mean state (immuno-competence) distribution so that the cumulative annual RTI count is matched to a reference situation (COPSAC2000 birth cohort[35]) (Figure S3). Note that, additionally, this procedure is done as a function of patient age with the immune effector function varying according to a sigmoid developmental dynamics model (Supplementary Methods: Impact of age).

#### Susceptibility of respiratory viruses to lower or upper tract

During the previously described Monte-Carlo-like process, the exposures were due to the main respiratory viruses - either RSV, rhinovirus or influenza. At each time point $t_i$, an exposure $ex_i$ is triggered and the corresponding viral trigger is chosen between the three viruses according to its current prevalence w.r.t. the seasonality. This means that the viral pathogen is chosen stochastically via the contribution of the specific seasonality of the virus, $p_{virus}(t_i)$ to the total seasonality of the three respiratory viruses $p(t_i)$ at the time point of exposure $t_i$. In a second step, if this exposure is converted to an RTI according to the combined deterministic-stochastic model, then we choose, again stochastically, whether an LRTI or URTI is provoked according to the viral affinity. We consider for each virus its unique probability to provoke a URTI conditioned on an RTI development (with the equivalent for LRTI stimulation being the complementary probability), which is used for this stochastic approach.

#### Calibration of OM-85 clinical efficacy

We performed the calibration of OM-85 clinical efficacy on top of the calibrated multi-scale RTI disease model. The unknown parameters left calibrated are those controlling the up-regulation of lymphocytes activation and virus-specific immunoglobulins production due to innate memory-like cells migrating from activated Peyer's Patches. Our main data source regarding OM-85 efficacy is the meta-analysis by Yin *et al.* (2018)[36] who performed a systematic review of 53 RCTs of its effect in recurrent RTI involving 4851 pediatric patients and reported frequency of RTIs in OM-85 vs. the control group. Their analysis showed that OM-85 was positively correlated with a reduction in the frequency of respiratory infection compared to the control group.

We grouped the studies into either 12 months or 6 months of follow-up and considered only studies with a single course of treatment and excluded all others. A 2D analysis of the absolute benefit as a function of the RTI frequency in the control group, similar to the Effect Model law[37] (Figure S9) confirms that the effect of OM-85 is in fact not a constant, but depends non-linearly on the risk for RTI (which explains that studies preferably enroll patients at risk for recurrent RTI). Selection of the appropriate at-risk population is usually done by evaluating the frequency of RTI in a reference period, assuming that immunological characteristics drive the risk for RTI and thus their frequency is correlated in consecutive years. This situation was mimicked by *in silico* clinical trials comprising an observational period and a follow-up period. We matched the simulated treatment efficacy distribution by small *in silico* clinical trials of 25 patients per arm with a pediatric population of 1 to 6 years of age while varying patients' eligibility criteria regarding their number of RTI in the observational period. The results of efficacy as a function of eligibility criteria (Figure S11-S10) indicate in fact that tuning the number of required RTI in the observational period can navigate through the entire meta-analyzed risk-stratified efficacy of the meta-analysis (parameters values controlling treatment efficacy were chosen from a series of simulations so that coverage the meta-analysis by the entire *in silico* procedure was maximal).

#### Mechanistic uncertainty management

To account for the substantial uncertainty regarding the mechanism determining the effect of OM-85 (through gut-reprogrammed innate cells that convey pro-type 1 immunomodulation in respiratory tract mucosal tissue), we simulated different mechanistic scenarios for the following key parameters in parallel as part of uncertainty management.

The first key but uncertain mechanism is the antigen presenting cell i.e. dendritic cell (DC) sensitivity to the concentration of the immunogenic compound, which depends on the administered dose. Indeed, human *in vivo* OM-85 dose-effect relationship data regarding the immune activation have not yet been reported. We used human *in vitro* data and apply an *in vitro* to *in vivo* translation factor. Since no data was available to set this factor, we included four different values for the DC sensitivity to OM-85 dose around a typical values of $1/10$[38].

Trained innate immunity postulates that immune cells can alter their gene expression for a longer time period after encountering an inflammatory stimulus and that they can partially persist in the organism and confer protection against a secondary stimulus[39,40]. Various *in vitro*, rodent models and clinical data suggest that exposure to OM-85 may generate generate such trained innate immune cells[41] enhancing a pre-alert anti-infectious RT-tissue state. This mechanism represented in our model through long-lived innate memory-like progenitor cells that can differentiate into type-1 innate immune cells. The lifetime of such postulated OM-85 stimulus-reprogrammed cells is unknown. We included three lifetimes: short (30 days), medium (60 days) and long (90 days).

The final model ensemble is then constructed with the Cartesian product of the set of values for the two key mechanisms and results are presented taking into account that uncertainty.

### Supplementary Figures and Tables

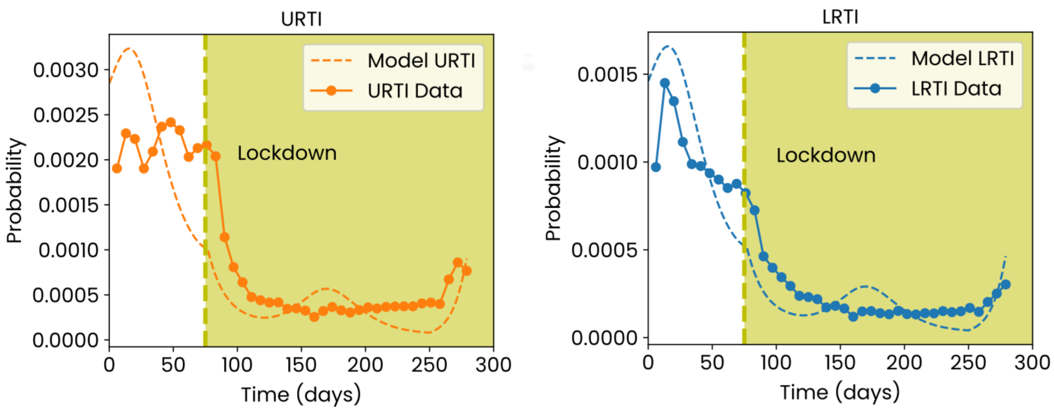


**Figure S1.** Validation of modelling approach of RTI burden under NPI measures using RCGP data[42]. Comparison between simulations and RCGP data on percentage of the population with either an upper or a lower RTI. The simulation starts at 01/01/2020, the lockdown is introduced at 75 days (mid-March) and is modelled by a transmission decreased by 17.5%.

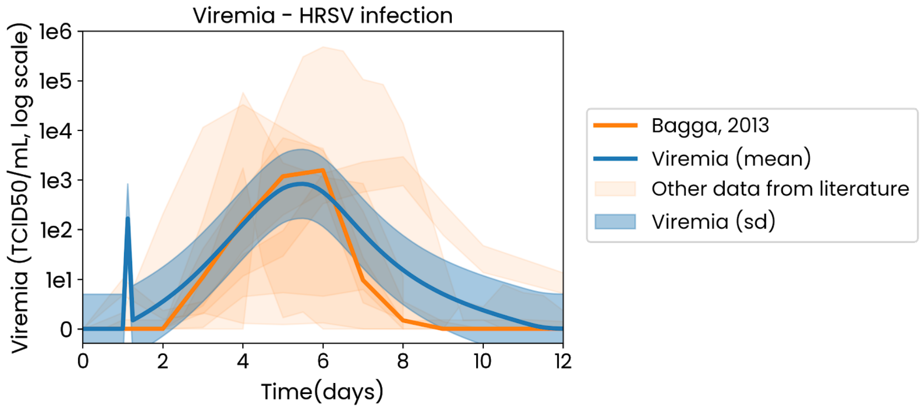


**Figure S2.** Viral load kinetics following experimental HRSV infection in adults. Experimental data (orange) was extracted from Bagga *et al.*[3]. Simulations are indicated in blue (mean and standard deviation). Virtual patients were challenged on day 1 with a viral dose of 1000 TCID50/mL. Other dataset from literature of experimental HRSV infections are superimposed (orange), confirming the general form of the viremia dynamics.

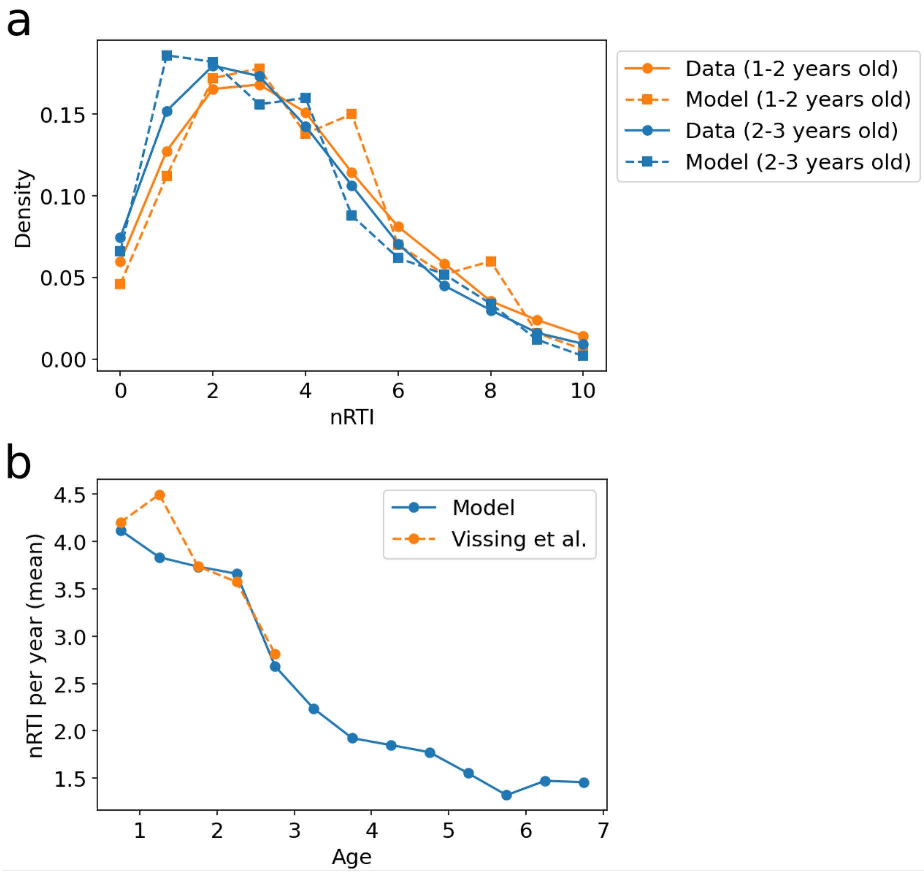


**Figure S3.** Calibration of immuno-maturation in the within-host model. **a**, Negative binomial distributions were fitted (solid lines) to match the distribution of number of RTIs (nRTI) from the COPSAC2000 cohort extracted from Vissing *et al.*[35] (median, mean and IQR for age 1-2 and 2-3 years old). Model parameters controlling immuno-maturation in the within-host RTI model were calibrated to reproduce this age-dependent distribution. Here a representative virtual cohort of similar size as the COPSAC2000 cohort (334 children) was simulated for two age groups (1-2 and 2-3 year old in orange and blue resp.) over one year. Distribution of cumulative number of RTIs for each age group is plotted (dashed lines). **b**, Number of RTIs per year per age is plotted for the model (solid blue line) for children from 0.5 to 7 years old and compared with data from the COPSAC2000 cohort[35].

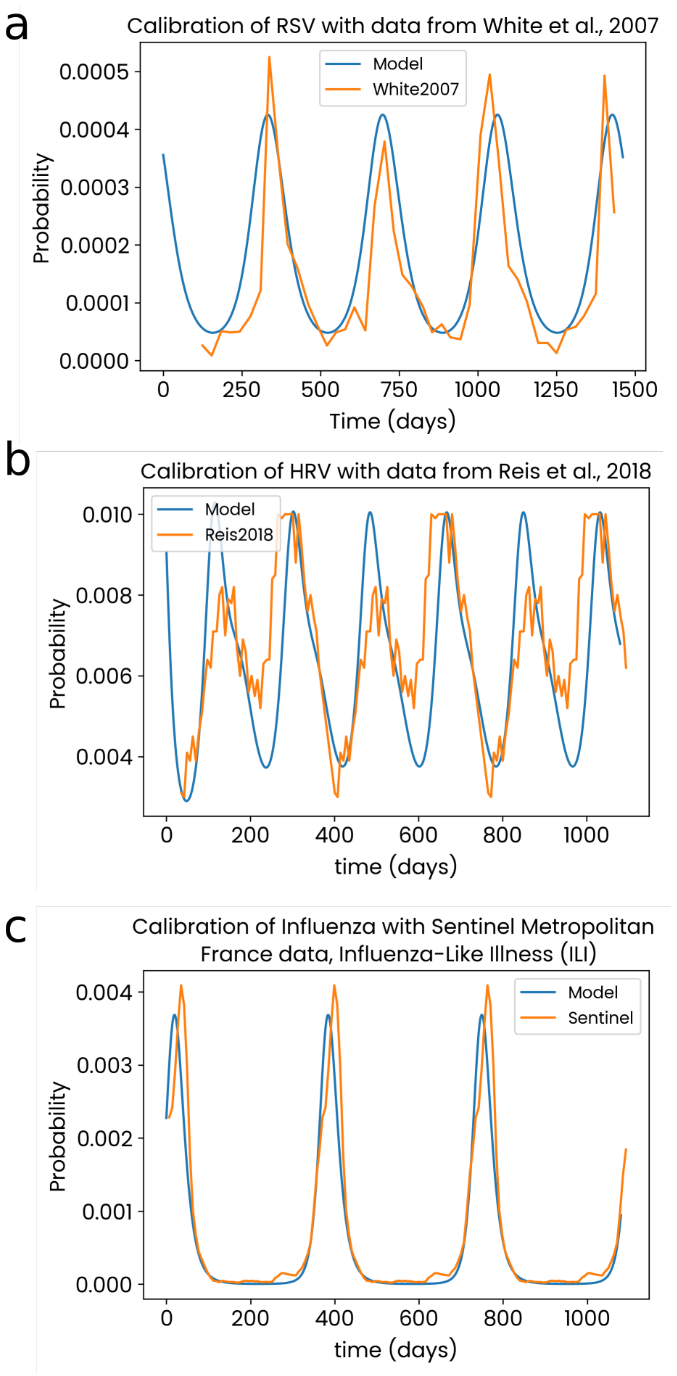


**Figure S4.** Calibration of the three parallel between-host SIRS models for a) respiratory syncytial viruses (RSV) with data from White *et al.* (2007)[11], b) rhinoviruses (HRV) with data from Reis *et al.* (2018)[43] and c) influenza viruses with data from Sentinel network in France[13]. Data over one year was replicated to extend the range of the comparison between simulations and data to several years when needed.

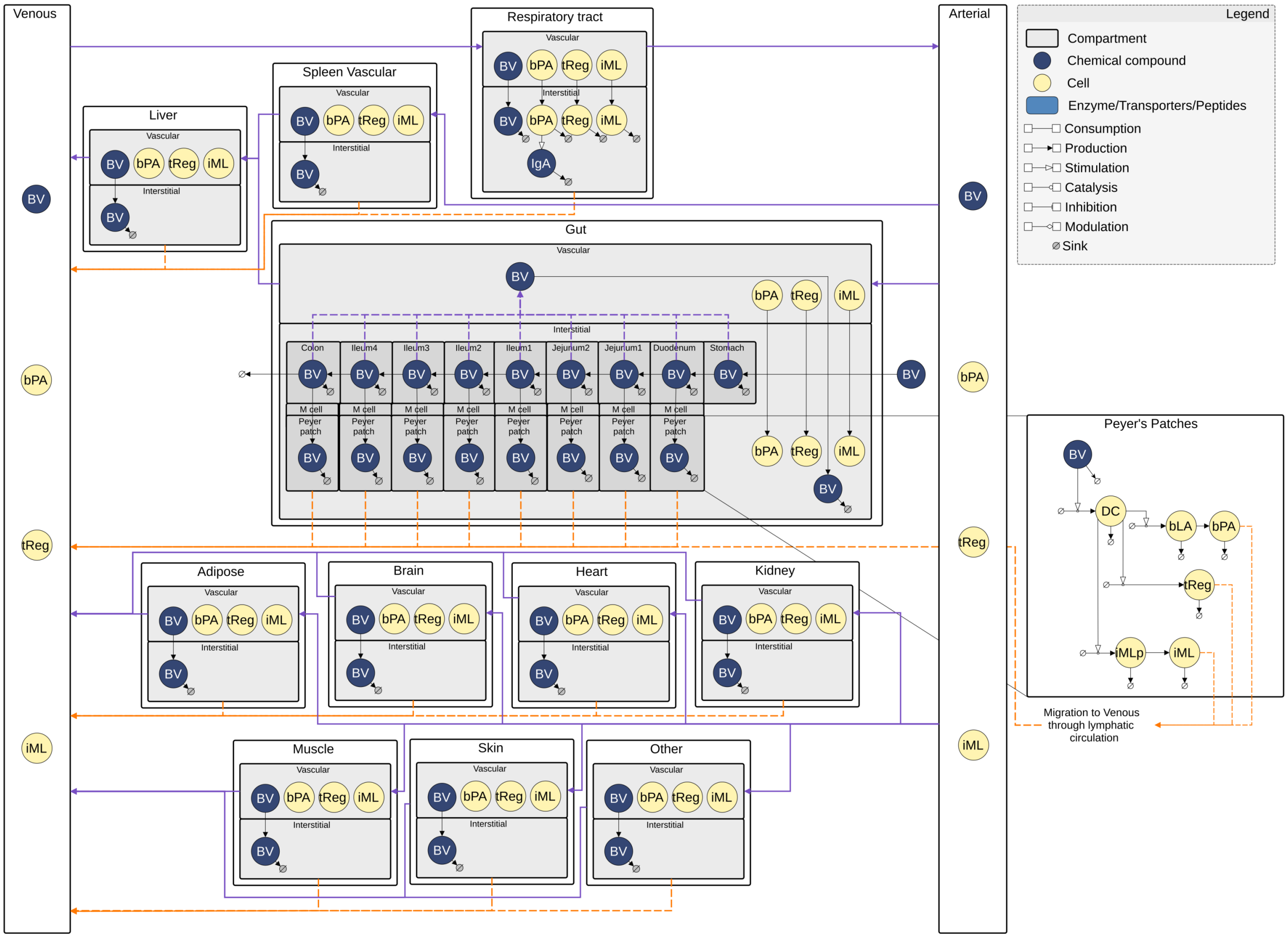


**Figure S5.** Graphical representation of PBPK/PD model used for the treatment model. A detailed absorption model is also represented notably for permeation via M cells to Peyer's Patches. BV = Broncho-Vaxom i.e. OM-85. The systemic circulation is indicated with purple arrows and the lymphatic circulation with orange arrows.

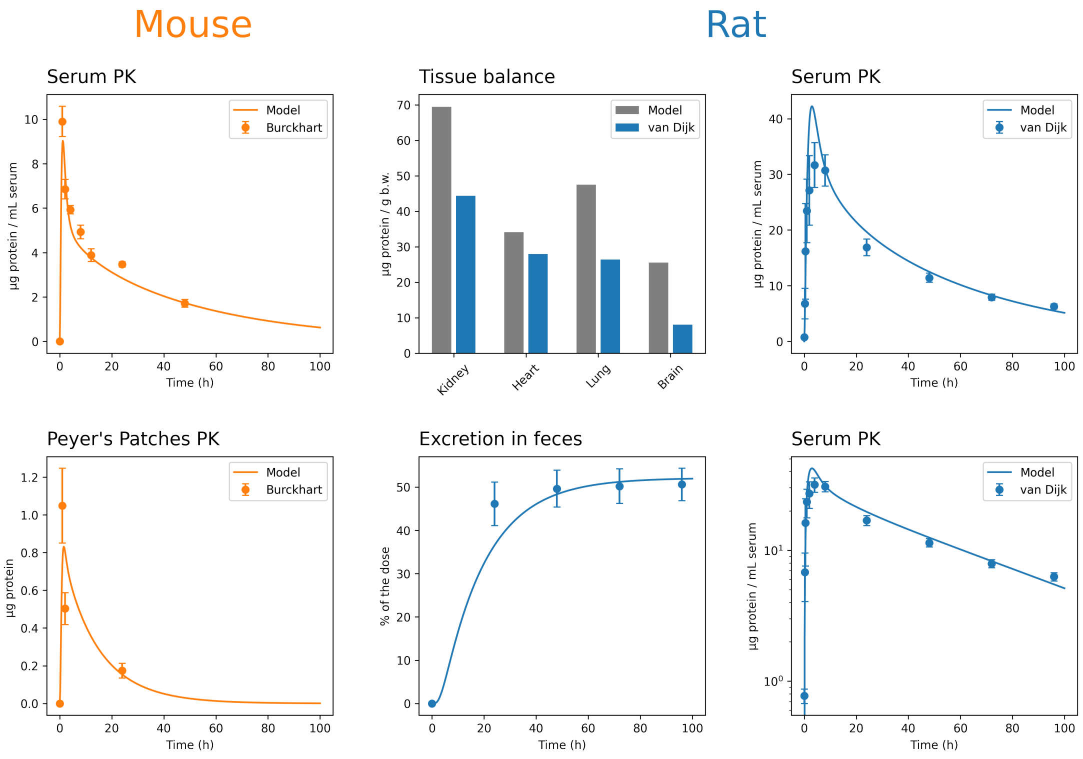


**Figure S6.** Calibration of the PBPK model with PK data on a radiolabelled derivative of a product (OM-89) similar to OM-85 in rodents[22,23]. We built a inter-species PBPK model by allometric scaling and species-specific parameters to use these data for calibration.

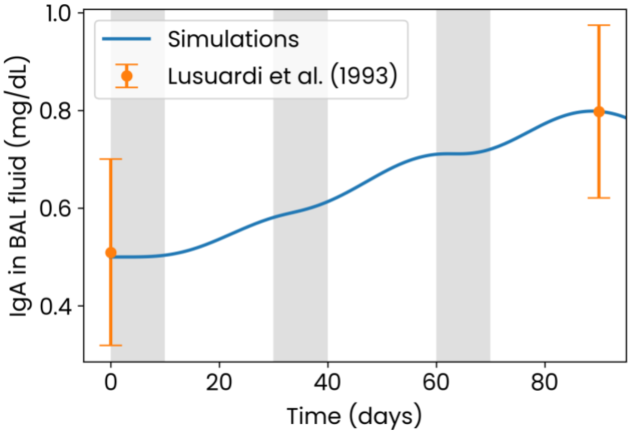


**Figure S7.** Final parametrization selected to reproduce the IgA dynamics in BAL fluid as reported by Lusuardi *et al.* (1993). Virtual patient is treated with three 10-day courses of OM-85 (light grey vertical bars).

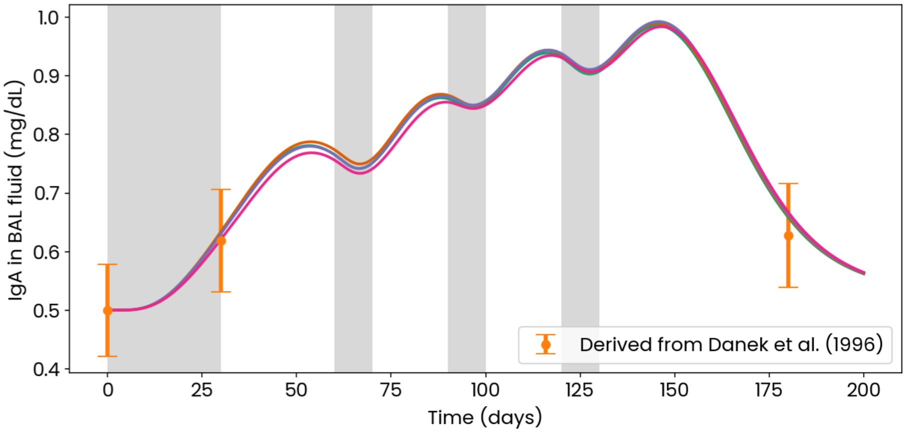


**Figure S8.** Validation data on IgA concentration in BAL fluid (mg/dL) derived from Danek et al. (1996)[32] vs simulations. Regimen: One month of daily administration followed by one month without treatment followed by 10 days of daily administration at the beginning of the month for 3 months. Phases of daily administration are marked in shaded grey. Note that simulations are reported here for the four different hypotheses on sensitivity to the dose of GALT sensor cells (professional antigen-presenting-cells) as described in the Methods.

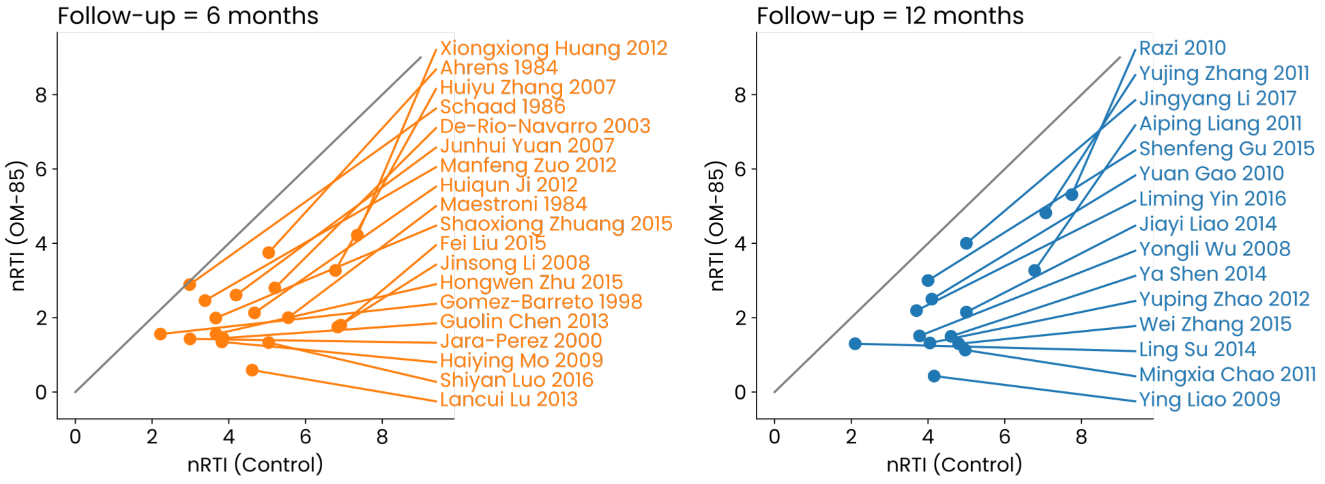


**Figure S9.** 2D analysis of OM-85's absolute benefit (number of prevented RTIs) as a function of the RTI frequency in the control group (similar to the Effect Model law[37]) using data from the meta-analysis by Yin *et al.* (2018)[36].

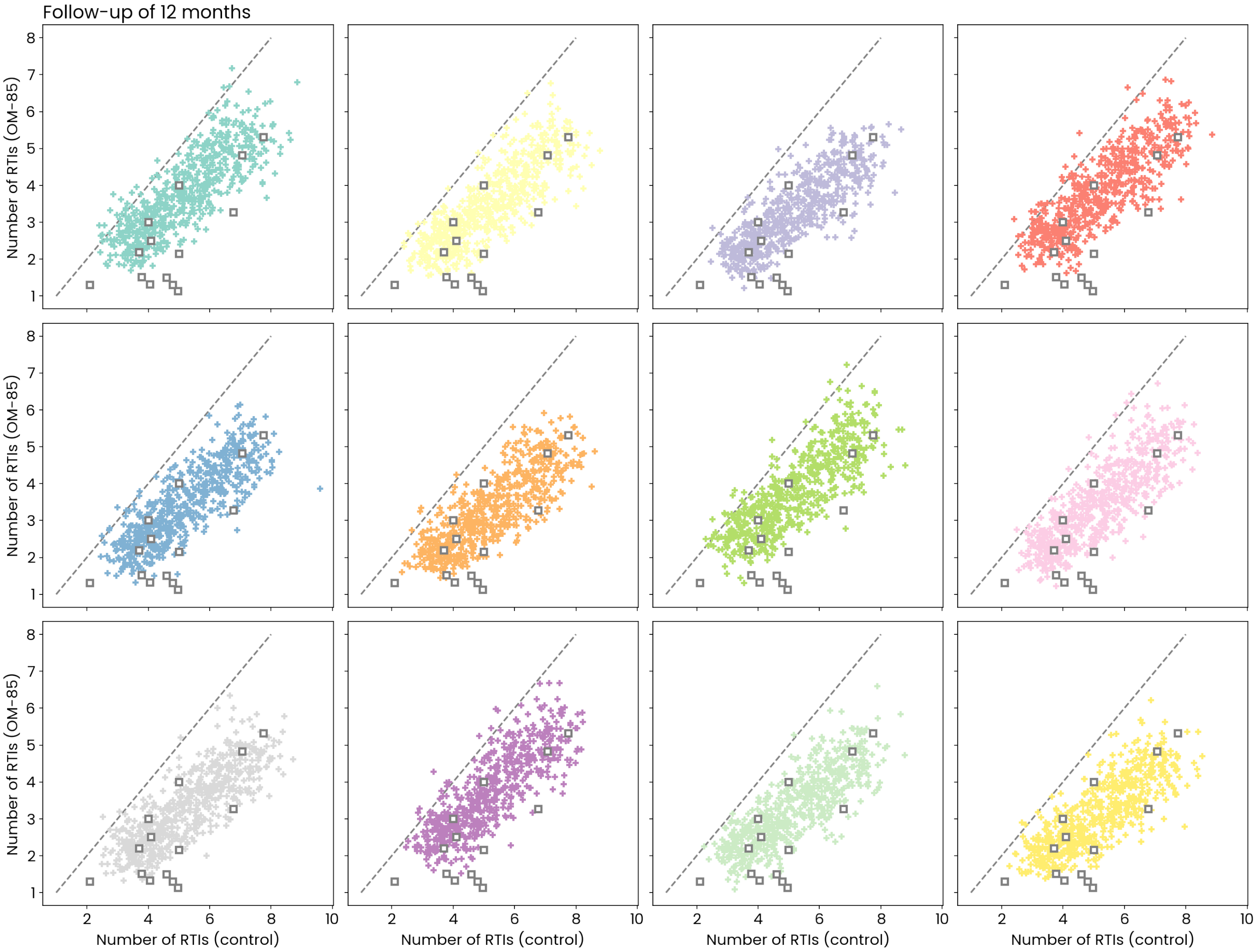


**Figure S10.** Comparison of OM-85's simulated efficacy with the data from the meta-analysis by Yin *et al.* (2018)[36] for a population of pediatric patients (1-6 years old) for a regular treatment of 10 daily administrations (3.5 mg) per month for 3 months followed by 12 months of follow-up. Here, for each model variation (defined by the colors, see Methods), we conducted 50 *in silico* trials with 50 patients per arm for various inclusion criteria (defined as the number of RTIs in the selection year prior to treatment) going from 0 up to 12 RTIs per year. For each of these trials, we report the mean number of RTIs during the follow-up period (12 months) in the control group vs the treated group (crosses). Points (squares) corresponding to real clinical trials obtained from the meta-analysis are overlayed.

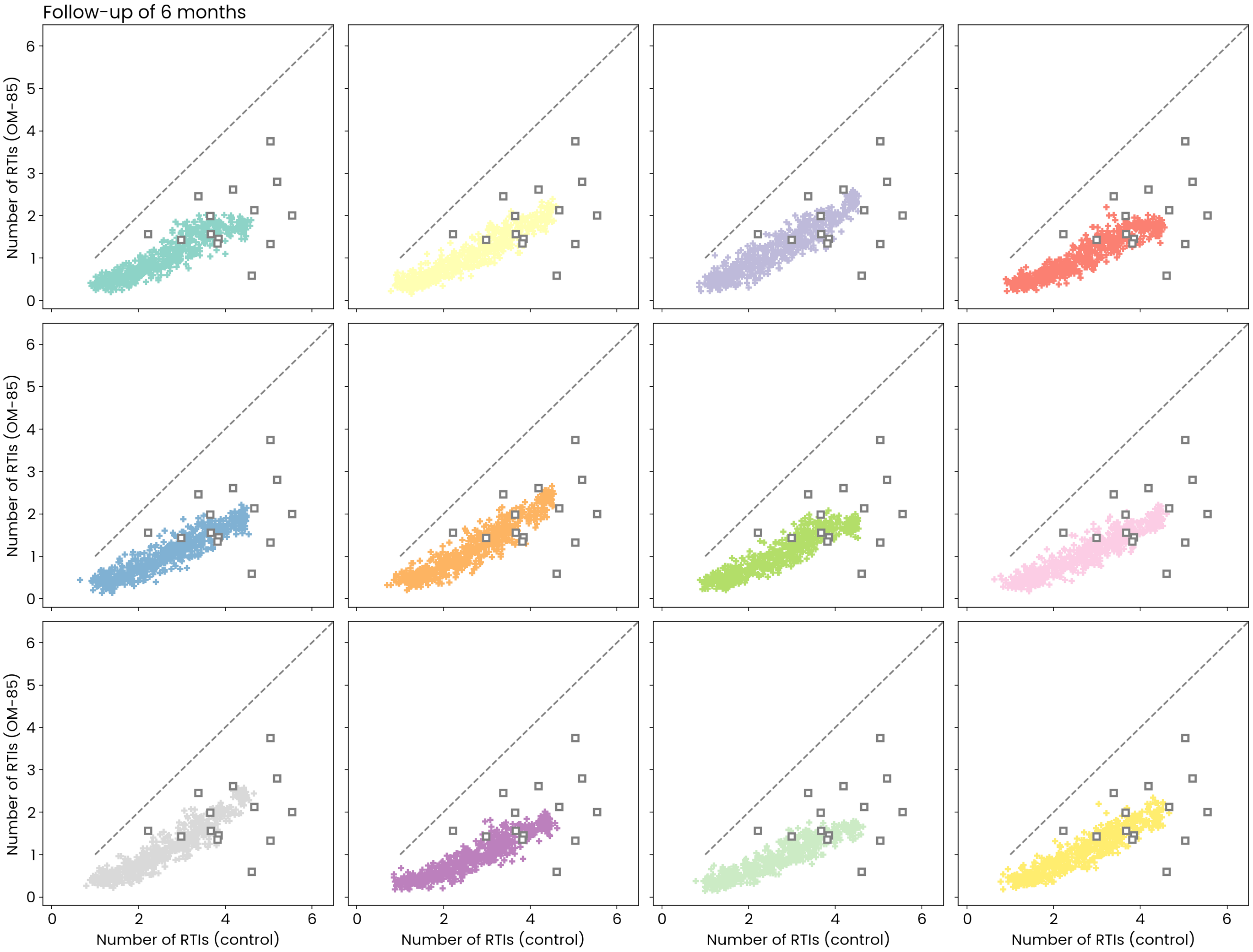


**Figure S11.** Comparison of OM-85's simulated efficacy with the data from the meta-analysis by Yin *et al.* (2018)[36] for a population of pediatric patients (1-6 years old) for a regular treatment of 10 daily administrations (3.5 mg) per month for 3 months followed by 6 months of follow-up. Here, for each model variation (defined by the colors, see Methods), we conducted 50 *in silico* trials with 50 patients per arm for various inclusion criteria (defined as the number of RTIs in the selection year prior to treatment) going from 0 up to 12 RTIs per year. For each of these trials, we report the mean number of RTIs during the follow-up period (6 months) in the control group vs the treated group (crosses). Points (squares) corresponding to real clinical trials obtained from the meta-analysis are overlayed.

**Table S1.** Parameters (description and value) of the within-host RTI disease model

| Parameter | Value | Unit | Description | Reference(s) |
|---|---|---|---|---|
| $n_i$ | 2.7 | $\mathrm{mL\,d^{-1}\,kcell^{-1}}$ | Lysis rate of infected cells by lymphocytes | Calibrated |
| $d_i$ | 0.26 | $\mathrm{d^{-1}}$ | Rate of infected cell death | Calibrated |
| $d_h$ | 0.01 | $\mathrm{s^{-1}}$ | Rate of healthy cell death | Calibrated |
| $p_h$ | 1 | $\mathrm{d^{-1}}$ | Epithelium renewal rate (percentage) | Calibrated |
| $d_v$ | $2 \cdot 10^{-6}$ | $\mathrm{d^{-1}}$ | Rate of viral particles decay | Calibrated |
| $k_{inf}$ | 0.11 | $\mathrm{mL\,d^{-1}\,kcell^{-1}}$ | rate of viral infection of healthy cells | Calibrated |
| $v_f$ | 1 | - | Virulence factor | Varied per virus type |
| $c$ | 10 | $\mathrm{count\,mL^{-1}}$ | Threshold under which the viral load will not affect the healthy cells (represents the preemptive clearance by the innate immune system) | Fixed |
| $p_c$ | $250 \cdot 10^6$ | $\mathrm{count\,d^{-1}}$ | Maximum production rate of pro-inflammatory cytokines | Calibrated |
| $K_c$ | $10 \cdot 10^6$ | $\mathrm{count\,mL^{-1}}$ | Modulation factor for the cytokine production | Calibrated |
| $d_c$ | 0.25 | $\mathrm{d^{-1}}$ | Natural decay rate of pro-inflammatory cytokines | Calibrated |
| $p_v$ | 2.58 | $\mathrm{d^{-1}}$ | Production rate of viruses by the infected cells | Calibrated |
| $p_{ig}$ | 3177 | $\mathrm{kcount\,mL^{-1}\,ms^{-1}}$ | Virus-specific antibody production | Calibrated |
| $d_{ig}$ | 0.22 | $\mathrm{d^{-1}}$ | Decay of virus-specific antibodies | Calibrated |
| $S$ | 3.78 | - | Immune state of the patient | Distributed in the population |
| $K_V$ | 608 | $\mathrm{kcount\,mL^{-1}}$ | Virus-mediated activation of the immune system | Calibrated |
| $p_M$ | 4 | - | Parameter upregulating the activation of the lymphocytes based on the innate memory-like cells resulting from GALT | Calibrated |
| $K_M$ | 4.64 | $\mathrm{cell\,mL^{-1}}$ | Modulates the activation of lymphocytes and IgA based on the innate memory-like cells resulting from GALT | Calibrated |
| $n_v$ | 10.7 | $\mathrm{\mu L\,d^{-1}\,Mcount^{-1}}$ | Neutralization rate of viruses by the antibodies | Calibrated |
| $p_L$ | 36.5 | $\mathrm{kcell\,mL^{-1}\,d^{-1}}$ | development rate of lymphocytes in response to infected cells | Calibrated |
| $d_L$ | 0.11 | $\mathrm{d^{-1}}$ | Death rate of the lymphocytes | Calibrated |
| $\bar{E}_h$ | 11.63 | $\mathrm{Mcell\,mL^{-1}}$ | Equilibrium concentration of healthy cells in the epithelium | Fixed |
| $d_{Ig_A}$ | 0.12 | $\mathrm{d^{-1}}$ | IgA decay rate | Fixed[44] |

**Table S2.** Parameters (description and value) of the between-host infection transmission model

| Parameter | Value | Unit | Description | References(s) |
|---|---|---|---|---|
| $\beta_{0,RSV}$ | 0.219 | $d^{-1}$ | mean value of infection rate $\beta$ for RSV | Calibrated |
| $\beta_{1,RSV}$ | 0.116 | - | relative magnitude of seasonal oscillations of $\beta$ for RSV | Calibrated |
| $\gamma_{RSV}$ | 0.1 | $d^{-1}$ | recovery rate for RSV | Fixed[9] |
| $\zeta_{RSV}$ | 0.005 | $d^{-1}$ | rate of loss of immunity for RSV | Fixed[9] |
| $P_{RSV}$ | 0.274 | - | phase shift of the $\beta$ seasonality as a fraction of a year for RSV | Calibrated |
| $\omega_{RSV}$ | $2\pi$ | - | frequency for seasonality oscillation of $\beta$ for RSV | Fixed |
| $\beta_{0,HRV}$ | 1.118 | $d^{-1}$ | mean value of infection rate $\beta$ for (human) rhinovirus | Calibrated |
| $\beta_{1,HRV}$ | 0.072 | - | relative magnitude of seasonal oscillations of $\beta$ for (human) rhinovirus | Calibrated |
| $\gamma_{HRV}$ | 0.5 | $d^{-1}$ | recovery rate for (human) rhinovirus | Fixed[45] |
| $\zeta_{HRV}$ | 0.0274 | $d^{-1}$ | rate of loss of immunity for (human) rhinovirus | Fixed[45] |
| $P_{HRV}$ | 0.596 | - | phase shift of the $\beta$ seasonality as a fraction of a year for (human) rhinovirus | Calibrated |
| $\omega_{HRV}$ | $4\pi$ | - | frequency for seasonality oscillation of $\beta$ for (human) rhinovirus | Fixed |
| $\beta_{0,IV}$ | 0.591 | $d^{-1}$ | mean value of infection rate $\beta$ for influenza | Calibrated |
| $\beta_{1,IV}$ | 0.101 | - | relative magnitude of seasonal oscillations of $\beta$ for influenza | Calibrated |
| $\gamma_{IV}$ | 0.2 | $d^{-1}$ | recovery rate for influenza | Fixed[46] |
| $\zeta_{IV}$ | 0.00274 | $d^{-1}$ | rate of loss of immunity for influenza | Fixed[46] |
| $P_{IV}$ | 0.036 | - | phase shift of the $\beta$ seasonality as a fraction of a year for influenza | Calibrated |
| $\omega_{IV}$ | $2\pi$ | - | frequency for seasonality oscillation of $\beta$ for influenza | Fixed |
| $A$ | 0.5992 | - | childcare influence | Calibrated |
| $L$ | 0.85 | - | effect of a lockdown scenario | Estimated via[42] |
| $f_{URTI}$ | 0.661 | - | fraction of URTIs in total number of RTIs | Estimated via[42] |
| $f_{LRTI}$ | 0.339 | - | fraction of LRTIs in total number of RTIs | Estimated via[42] |
| $N$ | 1 | - | population normalization (arbitrary number) | Fixed |
| $t_0$ | 1 | d | normalization time of the oscillations | Fixed |

**Table S3.** Drug-specific parameters (description and value) of the PBPK model. Parameters are given for mice and were allometrically scaled for rats and humans.

| Parameter | Value | Unit | Description |
|---|---|---|---|
| $\sigma_{PP}$ | 0.95 | - | Reflection coefficient in Peyer's Patches for immune cells |
| $\sigma_V^S$ | 0.15 | - | Scalar factor for vascular reflection coefficients |
| $p_{PP}^{Eff}$ | 34.4 | $\mathrm{nm\,s^{-1}}$ | Effective permeation of OM-85 into the PPs |
| $p^{Eff}$ | 376.4 | $\mathrm{nm\,s^{-1}}$ | Effective gut permeation of OM-85 |
| $d_{Gut}$ | 0.29 | $\mathrm{h^{-1}}$ | Degradation of OM-85 in the intestinal lumen |
| $CL$ | 8.4 | nL/ min /mg | Liver metabolic clearance of OM-85 per mg of microsomal proteins |
| $NSCL$ | 0.19 | μL/ min | Non-specific clearance of OM-85 per mg of microsomal proteins |

**Table S4.** Parameters of the pharmacodynamics model of OM-85 immune activation in Peyer's Patches.

| Parameter | Value | Unit | Description | Reference(s) |
|---|---|---|---|---|
| $E_O$ | 170 | $\mathrm{cell\,d^{-1}\,\mu L^{-1}}$ | Saturation factor for the activation rate of dendritic cell by OM-85 | Calibrated |
| $K_O$ | 0.12 | $\mathrm{\mu mol\,L^{-1}}$ | Half saturation constant for the activation rate of dendritic cell by OM-85 | Estimated from *in vitro* data[47] |
| $h$ | 3 | - | Hill coefficient for the activation rate of dendritic cell by OM-85 | Estimated from *in vitro* data[47] |
| $E_{M_p}$ | 550 | $\mathrm{cell\,d^{-1}\,L^{-1}}$ | Saturation factor for the activation rate of pre-activated type 1 innate cell by OM-85 activated DCs | Calibrated |
| $K_{M_p}$ | 200 | $\mathrm{cell\,\mu L^{-1}}$ | Half saturation constant for the activation rate of pre-activated type 1 innate progenitor cells by OM-85 activated DCs | Fixed |
| $E_{B_L}$ | 14 | $\mathrm{cell\,d^{-1}\,\mu L^{-1}}$ | Saturation factor for the activation rate of IgA+ B cells by OM-85 activated DCs | Calibrated |
| $K_{B_L}$ | 200 | $\mathrm{cell\,\mu L^{-1}}$ | Half saturation constant for the activation rate of IgA+ plasma cells by OM-85 activated DCs | Fixed |
| $E_{B_P}$ | 0.1 | $\mathrm{d^{-1}}$ | Differentiation rate of IgA+ B cells to plasma cells stimulated by OM-85 activated DCs | Calibrated |
| $K_{B_P}$ | 200 | $\mathrm{cell\,\mu L^{-1}}$ | Half saturation constant for the DC stimulated expansion of memory IgA+ B cells into IgA+ plasma cells | Fixed |
| $E_{T_r}$ | 10 | $\mathrm{cell\,d^{-1}\,\mu L^{-1}}$ | Saturation factor for the rate of activation of tRegs by OM-85 activated DCs | Calibrated |
| $K_{T_r}$ | 200 | $\mathrm{cell\,\mu L^{-1}}$ | Half saturation constant for the rate of tRegs by OM-85 activated dCs | Fixed |
| $\alpha$ | 0.5 | $\mathrm{d^{-1}}$ | Differentiation rate of reprogrammed type-1 innate progenitors in innate memory-like cells | Fixed |
| $\beta$ | 0.01 | $\mathrm{d^{-1}}$ | Basal differentiation rate of IgA+ B cells to plasmacells | Fixed |
| $\gamma$ | 200 | $\mathrm{pg\,cell^{-1}\,d^{-1}}$ | Non-specific IgA production rate by plasma cells | Fixed[48,49] |
| $p_{I_{gA}}$ | 0.06 | $\mathrm{mg\,d^{-1}\,dL^{-1}}$ | IgA basal production rate | Fixed[50] |
| $d_D$ | 0.11 | $\mathrm{d^{-1}}$ | Death rate of OM-85 activated dendritic cell | Fixed[51] |
| $d_{M_p}$ | 0.01 | $\mathrm{d^{-1}}$ | Death rate of pre-activated type 1 innate cell | Calibrated |
| $d_M$ | 0.06 | $\mathrm{d^{-1}}$ | Natural decay rate of pre-activated type 1 innate cell | Calibrated |
| $d_{B_L}$ | 0.005 | $\mathrm{d^{-1}}$ | Death rate of IgA+ B cells | Fixed[52] |
| $d_{B_P}$ | 0.23 | $\mathrm{d^{-1}}$ | Death rate of non-specific IgA+ plasmacells | Fixed[49] |
| $d_{T_r}$ | 0.12 | $\mathrm{d^{-1}}$ | Death rate of regulatory T cells | Fixed[52] |
| $d_{I_{gA}}$ | 0.12 | $\mathrm{d^{-1}}$ | IgA decay rate | Fixed[44] |
